# Distribution of Vector Abundance and Infection Rates in Relation to Human Vector-Borne Disease Cases in Nebraska

**DOI:** 10.64898/2026.02.12.26346200

**Authors:** Sarah A. Uhm, Halie Smith, Su Chen, Peter C. Iwen, Emily L. McCutchen, Amanda M. Bartling, Roberto Cortiñas, David Brett-Major, M. Jana Broadhurst, Jeff Hamik, Joseph R. Fauver

## Abstract

Vector-borne diseases represent a growing public health issue nationwide. Nebraska reports a sustained burden of mosquito-borne diseases and expanding tick-borne disease risk. This study aims to assess trends in vector abundance, vector infection rates, and human vector-borne disease reports using retrospective surveillance data and to examine the relationship between vector factors and human risk across the state. Vector abundance and pathogen infection rates were mapped alongside presence and incidence of key vector-borne diseases. Mosquito surveillance and mosquito-borne disease data were available from 2012-2024, while tick surveillance and associated tick-borne disease data were available from 2021-2024. Statistical models and comparative tests were used to explore associations between vector factors and disease reports. Overall, the current vector-borne disease surveillance suggests that mosquito-borne disease remains the primary concern in Nebraska, with notable geographic variation in mosquito species distribution and human cases. Tick surveillance indicates established populations of clinically relevant tick species in distinct regions of the state, with pathogen detections generally aligning with areas where human cases have been reported. Differences in the level of human case reporting and data availability affected interpretation of long-term trends and limited strong conclusions regarding direct relationships between vector factors and human disease. Nebraska maintains comprehensive vector-borne disease surveillance systems that support understandings of vector abundance, vector pathogen infection rates, and human risk. Continued integration of entomological surveillance with human case information may help clarify patterns of vector-borne disease risk and inform efforts to address current and emerging threats in Nebraska.

## Introduction

Vector-borne diseases (VBDs) pose a significant risk to public health, contributing to increased human morbidity, mortality, long-term disabilities, and strain on healthcare systems (Beard et al. 2019). Globally, VBDs comprise >17% of all infectious diseases, leading to hundreds of thousands of deaths annually (WHO 2024). Between 2001 and 2023, over 1 million VBD cases were reported in the United States, with cases doubling over the past 20 years as vector ranges expand and human exposures increase (CDC 2024a, 2024c). These VBD cases also contribute to severe outcomes such as neuroinvasive disease, chronic illness, pregnancy complications, and death (CDC 2024a, CDC 2024c, CDC 2024d, Sejvar 2014). West Nile virus (WNV) infection is the leading cause of mosquito-borne diseases (MBDs) in the United States, while Lyme disease is the leading cause of tick-borne diseases (TBDs) (CDC 2024d, Sutter et al. 2024). However, numerous other VBDs such as anaplasmosis, dengue fever, Zika virus infection, Powassan virus infection, and many others, have also been observed throughout the country (CDC 2024a, 2024c). Beyond the public health burden, VBDs also impose significant economic costs. Diagnosed Lyme disease cases cost upwards of $345 million USD every year while the annual costs of WNV infection are around $56 million USD, with estimates increasing considerably when patients are hospitalized with neuroinvasive disease (Barrett 2014, Staples et al. 2014, Hook et al. 2022). The expanding public health burden and economic impact of VBDs underscore the critical need for comprehensive surveillance, prevention, and early detection strategies.

VBDs represent a substantial and growing health burden throughout the Great Plains region of the United States, and Nebraska in particular. This area has the highest cumulative incidence of WNV infection in the United States (Karim and Bai 2023, Brüssow and Figuerola 2025). Since the introduction of WNV into the United States in 1999, this virus has become endemic, with population serosurveys showing that up to 20% of people in western Nebraska show IgG to WNV (Schweitzer et al. 2006, Carson et al. 2012). In the last five years, Nebraska has reported over 400 human cases to the Centers for Disease Control and Prevention (CDC), more than 200 of which were cases of neuroinvasive disease (Smith et al. 2020, CDC 2024b). However, up to 80% of WNV infections are subclinical and therefore less likely to be tested for and reported, masking the burden of WNV disease (Sejvar 2014). Although less than 1% of WNV infections lead to neuroinvasive disease, the severe manifestations of meningitis, encephalitis, and acute flaccid myelitis, when neuro-invasion does occur, are severe and can often lead to lasting neurological deficits (Sejvar 2014). Additionally, even mild illness can cause long-term neurological sequelae, underscoring the significant impact of WNV (Fulton et al. 2020). TBDs are historically less common in Nebraska in comparison to the Northeast and Upper Midwest United States (Boulanger et al. 2025). However, in recent years an increased number of reported TBD cases, including ehrlichiosis, rickettsiosis, and Lyme disease have been reported in Nebraska (NDHHS 2024a). The TBD landscape is also fast-changing, as evidenced by the first detection of *I. scapularis* and subsequent transmission of Lyme disease in the northeastern region of the state (Nielsen et al. 2020, Hamik et al. 2023). Although not involving a pathogen but caused by tick bites, the allergy to alpha-gal in mammalian products, known as alpha-gal syndrome (AGS), recently became a reportable condition in Nebraska, with case monitoring beginning in January 2025 (NDHHS, 2025a). As of October 2025, there have been 24 cases of alpha-gal syndrome in Nebraska (NDHHS, 2025b). Additionally, while not yet detected in Nebraska, a confirmed case of theileriosis caused by the protozoan parasite, *Theileria orientalis*, which is transmitted by the Asian longhorned tick, was identified in Iowa cattle in June 2025, demonstrating the persistent threat of novel TBD introduction in the state (Kaisand 2025).

To combat the threat posed by VBDs in the state, the Nebraska Department of Health and Human Services (DHHS) works with local health departments to conduct annual entomological surveillance for both mosquito- and tick-borne diseases. To collect mosquitoes, traps are set in key locations in counties throughout the state every other week from late May/early June to the end of September or the first hard freeze. The collected mosquitoes are then tested by the Nebraska Public Health Laboratory (NPHL) for the presence of WNV, St. Louis encephalitis virus (SLEV), and western equine encephalitis virus (WEEV). Mosquito abundance and pooled infection rates are calculated at the county scale to inform contemporary MBD risk. Data from Nebraska and neighboring states highlight *Culex tarsalis* mosquitoes as an important WNV vector in the Great Plains region, however, *Culex pipiens* and other related species may also contribute to WNV transmission in more urban environments (Schweitzer et al. 2006, Dunphy et al. 2019). It is currently unclear how each species contributes to WNV transmission, as is the quantitative association between vector abundance and infection rates and human WNV disease incidence in Nebraska. Active tick surveillance in Nebraska involves collecting questing ticks by dragging and flagging vegetation. Surveillance efforts have demonstrated *Dermacentor variabilis* as the most common tick species collected in the state, followed by *A. americanum* and *I. scapularis* at lower numbers (NDHHS 2024b). Ticks are then tested either in pools or as individuals for the presence of tick-borne bacteria and viruses, including the causative agents of Lyme disease, ehrlichiosis, rickettsiosis, anaplasmosis, tularemia, and more. It remains to be determined how infection rates in vector tick species are associated with occurrence of human cases of TBD in Nebraska.

In this study, retrospective entomological surveillance data and human case data in Nebraska were used to assess trends in vector abundance, vector infection rates, and human cases of both mosquito- and tick-borne diseases. We aimed to determine the association of vector prevalence and infection rates with diagnosed human disease cases. Additionally, we assessed whether variation in mosquito and tick infection rates correlates with spatial patterns of human VBD incidence across Nebraska. By linking entomological and epidemiological data, we assessed whether infection rates in vector populations provide actionable indicators of human risk.

## Materials and Methods

Entomological and epidemiological data from multiple sources were compiled to assess the relationship between vector infection rates and human VBD risk in Nebraska. Publicly available United States Census county-level estimates were used for Nebraska county populations.

### Data Sources and Approach: Overview of Vector-Borne Diseases in Nebraska

Nebraska DHHS provided publicly available data from 2010-2024 on MBDs and TBDs in the state. To visualize the general trends in this data, we applied several complementary analyses to link our vector surveillance and pooled vector infection rate data with human VBD cases. We graphed the total numbers of MBDs and TBDs by year and reported the total numbers of confirmed cases of anaplasmosis, ehrlichiosis, Lyme disease, rickettsiosis, tularemia, and neuroinvasive and non-neuroinvasive WNV (Figure 1 and Table 1).

**Table 1.** Total diagnosed cases of Vector-Borne Disease in Nebraska. Total reported human cases of vector-borne diseases from 2010-2024 in Nebraska, separated by specific tick-borne and mosquito-borne diseases.

| Vector-Borne Disease | Number of Reported Human Cases |
| --- | --- |
| Tick-Borne |  |
| Anaplasmosis | 12 |
| Ehrlichiosis | 105 |
| Lyme Disease | 145 |
| Rickettsiosis | 233 |
| Tularemia | 170 |
| Mosquito-Borne |  |
| WNV (Neuroinvasive) | 631 |
| WNV (Non-Neuroinvasive) | 935 |

**Figure 1.**
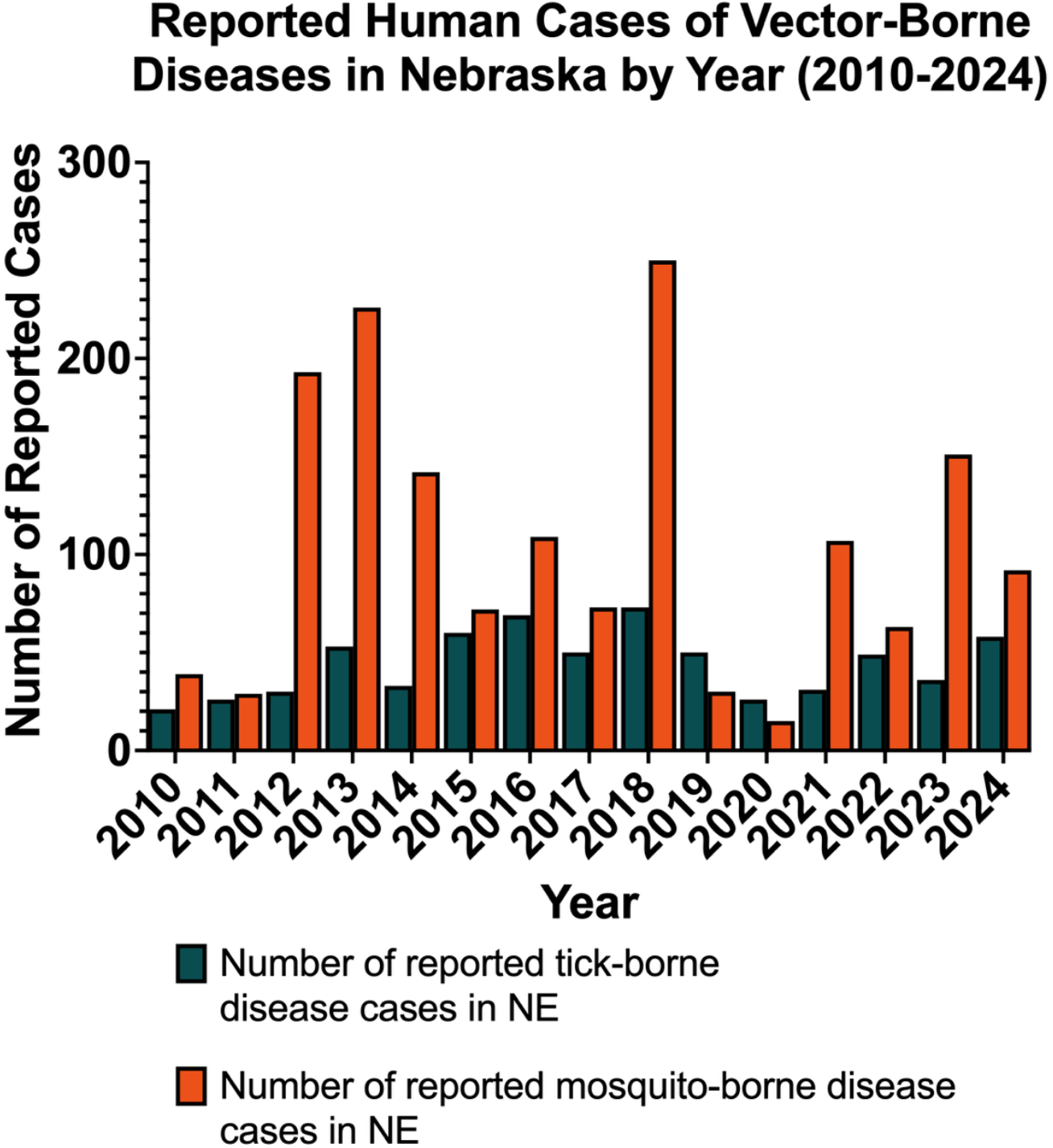
Cases of vector-borne diseases in Nebraska by year. Reported human cases of vector-borne diseases by year from 2010-2024 in Nebraska, separated by tick-borne and mosquito-borne cases.

### Mosquito Surveillance and Mosquito-Borne Diseases

CDC ArboNET provided human case data and demographic information for MBDs, specifically WNV infection, from 2012-2024 at the county level. While other arboviruses likely circulate in the state (e.g. St. Louis encephalitis virus), no clinical cases were reported during the study period. Additionally, arboviruses that were likely imported (i.e. not autochthonous), including Zika, dengue, and chikungunya viruses were excluded from analysis. Nebraska DHHS and the NPHL provided data from ongoing mosquito collections from light traps placed in counties throughout the state and pooled testing for WNV from 2012-2024.

For WNV, the average annual cumulative incidence of human cases per 100,000, combining neuroinvasive and non-neuroinvasive cases was determined for the 82 out of 93 Nebraska counties that reported cases to the CDC from 2012-2024. The most up to date 2024 county population estimates were defined as the total population at risk for the cumulative incidence calculations (U.S. Census Bureau, 2025).

For the 40 counties where Nebraska DHHS conducted mosquito trapping from 2012-2024, the average annual abundance per trap night for *Cx. tarsalis* and *Cx. pipiens* was calculated. Utilizing the Centers for Disease Control and Prevention’s (CDC, 2024) PooledInfRate R Studio package, WNV pooled infection rates per 1,000 mosquitoes for *Cx. tarsalis* and *Cx. pipiens* were determined (CDC 2024e).

The average annual WNV human case cumulative incidence per 100,000 people, the average annual abundance per trap night by mosquito species, and the average annual mosquito WNV pooled infection rates per 1,000 mosquitoes were calculated to visualize the overall trends across Nebraska. Representative state maps were created in ArcGIS Pro, comparing *Cx. tarsalis* and *Cx. pipiens* average annual abundance and pooled infection rates with WNV human case cumulative incidence at the county level (Figure 2). Spatial autocorrelation (Global Moran’s I) was used to evaluate the clustering pattern of WNV human cases. Our goal was to assess the overall long-term trends in this retrospective data, not to assess year to year differences.

**Figure 2.**
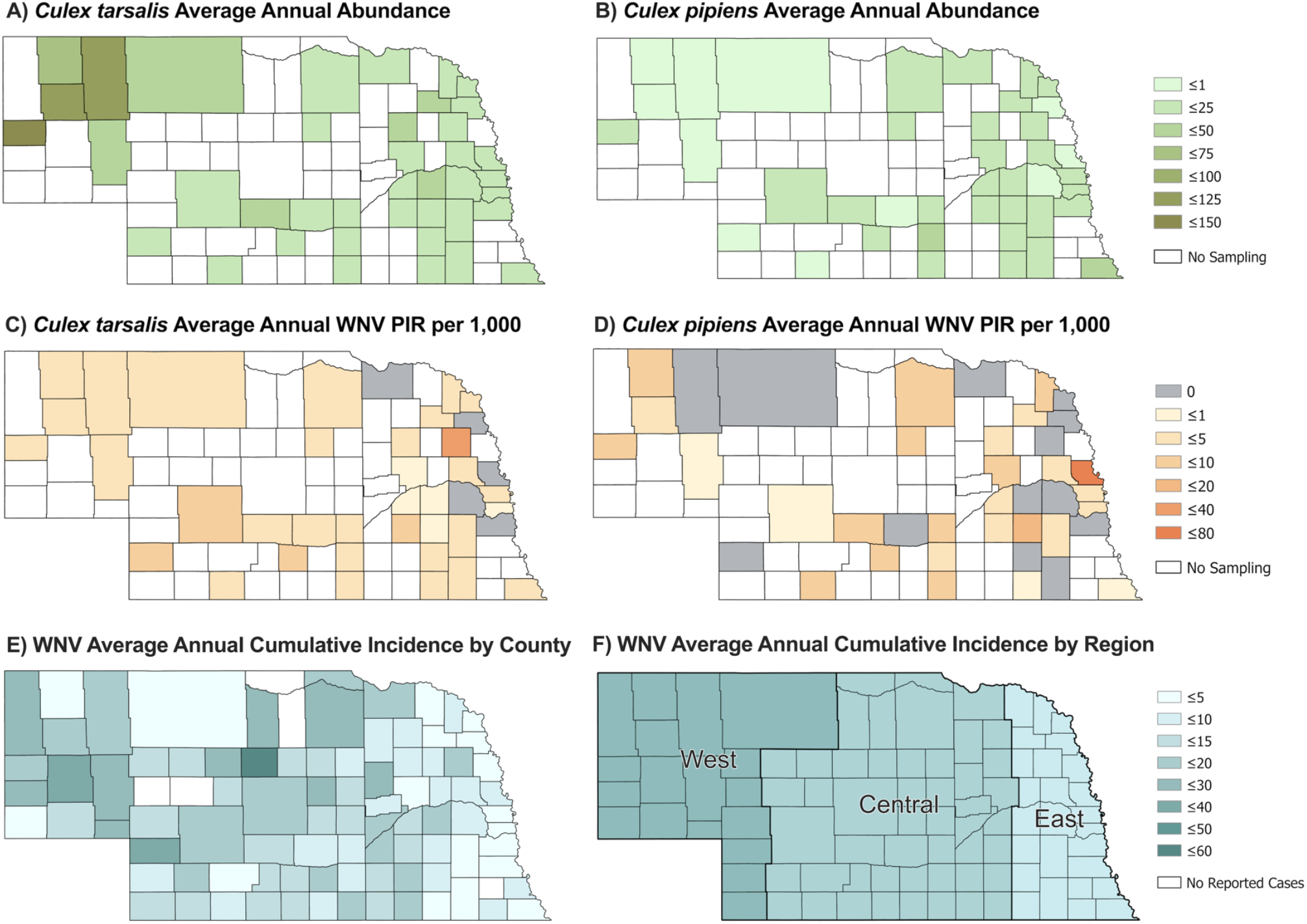
Comparison of *Culex* species abundance, pooled infection rate, and human WNV cases. A) *Culex tarsalis* average annual abundance per trap night and B) *Culex pipiens* average annual abundance per trap night (right) from 2012-2024. C) *Culex tarsalis* average annual WNV pooled infection rate per 1,000 mosquitoes and D) *Culex pipiens* average annual WNV pooled infection rate per 1,000 mosquitoes from 2012-2024. E) WNV average annual cumulative incidence per 100,000 people from 2012-2024 by county and F) WNV average annual incidence per 100,000 people from 2012-2024 by region, highlighting concentration of cases in western Nebraska.

To assess the relationships between *Culex* species-specific annual abundance per trap night or annual pooled infection rates and WNV human cases from 2012-2024, we assessed the normality of the data (Shapiro-Wilk test) and created scatterplots with line fits and LOESS (locally estimated scatterplot smoothing) between each variable (*Culex* species-specific data) and outcome (WNV human cases). To assess the data for heteroscedasticity, we used a Breusch-Pagan test, which fits a linear regression model to the residuals of a linear regression model. A Cameron-Trivedi score test to detect any overdispersion in our WNV human case count data was also performed. A negative binomial model was determined to be the best fit for the outcome of WNV human case counts with *Cx. tarsalis* abundance, *Cx. tarsalis* WNV pooled infection rate, *Cx. pipiens* abundance, and *Cx. pipiens* pooled infection rate as predictor variables.

To achieve a more contemporary perspective on the relationships between *Culex* species-specific annual abundance per trap night or annual pooled infection rates and WNV human cases, we focused on Nebraska counties reporting both mosquito and WNV human case data for at least three of the four years from 2021-2024. With this new data subset of nine counties, we reran our tests for normality (Shapiro-Wilk test) and created scatterplots with line fits and LOESS curves were rerun. This data subset was also assessed for heteroscedasticity while a Cameron-Trivedi score test was used to detect overdispersion.

We evaluated the underlying distributions of this contemporary data subset with total WNV case counts and cumulative incidence per 100,000 using maximum-likelihood fitting. For each outcome (WNV case counts and cumulative incidence per 100,000), multiple candidate probability families appropriate to data type (e.g., Poisson, negative binomial, lognormal, and gamma) were tested. Model performance was compared using Akaike Information Criterion (AIC) and Bayesian Information Criterion (BIC), and the best-supported distribution was selected for further use. Goodness-of-fit was visualized using density, CDF, QQ, and PP comparisons.

For this contemporary data subset, a negative binomial model with log link was used to model the predictor variables in relation to the count outcome of WNV human cases. A gamma with log link model was used to model predictor variables in relation to the outcome of cumulative incidence per 100,000. Because the dataset included only nine counties and four years, both county and year were modeled as fixed effects to allow for an estimation of county-specific baseline differences and account for known spatial clustering of human WNV cases in the western part of the state, while adjusting for statewide interannual variation.

### Tick Surveillance and Tick-Borne Diseases

Human case data for tick-borne diseases from 2021-2024 at the health department level from the Nebraska Department of Health and Human Services (DHHS) was obtained. For tick surveillance information, Nebraska DHHS shared species abundance and pathogen testing from 2021-2024. TBD human case data were reported at the health department level and any counts less than six were listed as “suppressed”. The analysis of TBD cases was subsequently approached at the health department level, rather than the county level, with cases of TBDs organized by the presence or absence of reported cases. However, as tick surveillance was conducted at the county level, the tick surveillance data were extrapolated to the health department level to compare entomological information with human cases.

For the 56 counties where Nebraska DHHS conducted tick dragging and flagging from 2021-2024, the average annual abundance per square meter for *A. americanum, D. variabilis*, and *I. scapularis* were calculated. This tick abundance data was subsequently visualized by creating representative maps in ArcGIS Pro. Tick abundance averages were visually compared to publicly available information from Nebraska DHHS regarding tick population distribution and population status (established, reported, or no known populations) at the county level (Figure 4). CDC definitions of established and reported were used in these determinations (Dennis et al. 1998). The CDC PooledInfRate R Studio package was used to determine the pooled infection rates per 1,000 for *A. americanum, D. variabilis*, and *I. scapularis* by each county, year, and reported tick-borne pathogen that causes disease in humans. The average annual pooled infection rates per 1,000 were then determined for each tick species by county, year, and pathogen across the years 2021-2024 (Table 2). Infection rates were calculated and compared in species of ticks that were tested for the presence of a specific pathogen. The number of years (from 2021-2024) with confirmed human cases of anaplasmosis, ehrlichiosis, rickettsiosis, and tularemia at the health department level were also visualized with ArcGIS Pro (Figure 5). Lyme disease was not included in the primary analyses as autochthonous transmission is currently established only in limited areas of the northeastern region of the state.

**Table 2.**
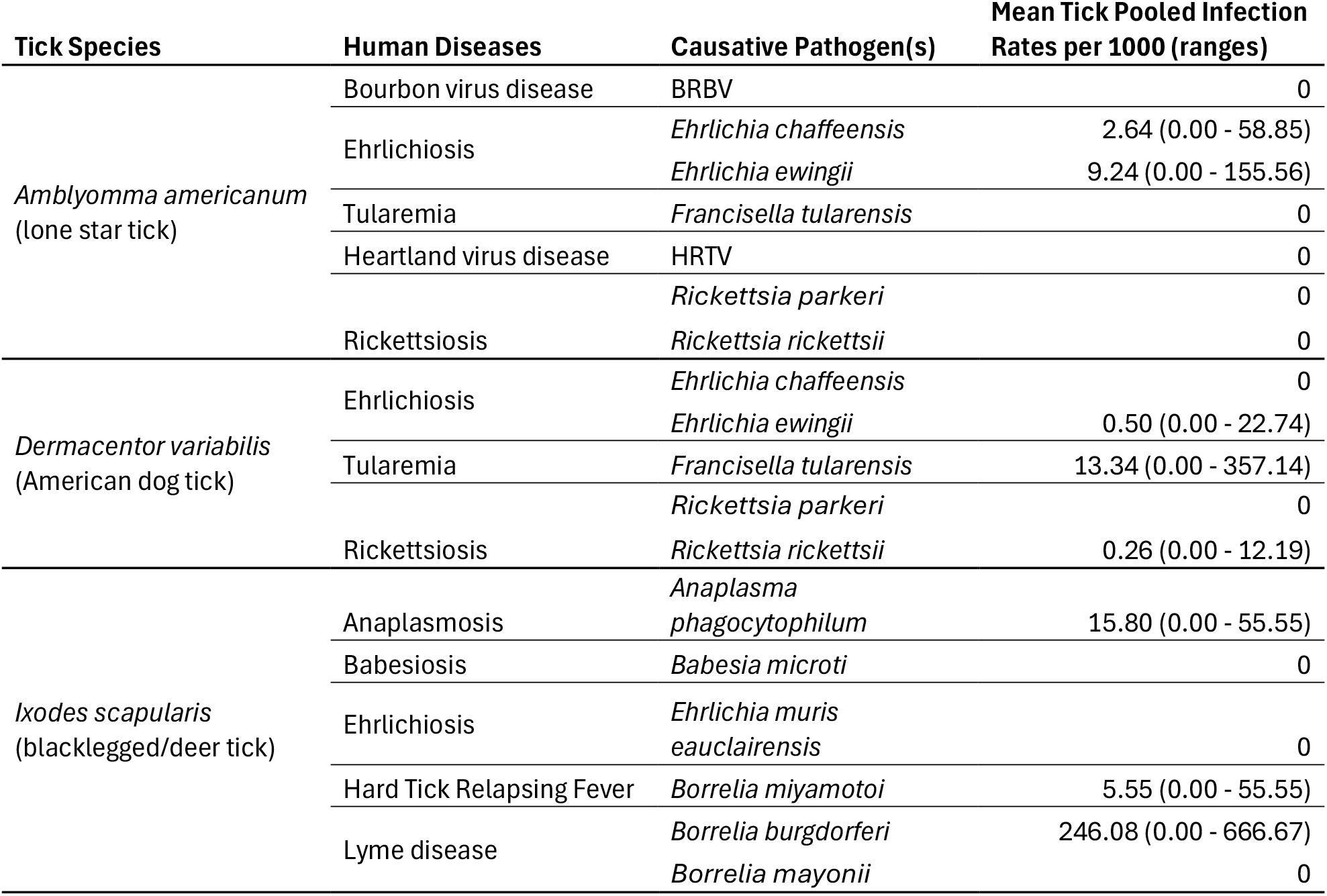
Overview of tick pooled infection rates by species and pathogen. Tick species, human disease, causative pathogen vectored, and summary statistics for the pooled infection rates per 1,000 ticks for each pathogen from 2021-2024 in Nebraska.

To assess the relationship between tick pooled infection rates and human case incidence of TBDs, we focused on anaplasmosis, ehrlichiosis, rickettsiosis, and tularemia. Initially, logistic regressions for each of these four TBDs were conducted using a generalized linear model and a generalized linear mixed-effects model, which indicated no significant relationships. Considering that limited cases (e.g. few health departments reported human cases) can potentially lower logistic regression power, a Wilcoxon rank-sum test for each of the four TBDs was used to determine if there were any significant differences in the tick pooled infection rates between health departments that reported no human cases of a given disease compared to departments that reported confirmed cases (Figure 6).

## Results

### Overview of Vector-Borne Diseases in Nebraska

From 2010 to 2024, Nebraska consistently reported cases of both MBDs and TBDs annually. However, the prevalence of MBD and TBD cases showed noticeable variability over time with distinct peaks in 2012, 2013, 2018, and 2023 (Figure 1). Overall, MBDs comprised the majority of VBD cases across the state. Of the MBDs, WNV infection was the primary disease reported (Table 1). TBDs were less common than MBDs. Lyme disease, rickettsiosis, and tularemia were the most commonly reported TBDs during the study period.

### Mosquito Surveillance: Cx. tarsalis and Cx. pipiens Abundance and WNV Pooled Infection Rates

The average annual *Cx. tarsalis* and *Cx. pipiens* abundance per trap night from 2012-2024 revealed consistent differences in the spatial distribution of these species (Figure 2A and 2B). *Culex tarsalis* abundance was highest in western Nebraska, while *Cx. pipiens* was observed more in the eastern regions. The average annual pooled infection rates (PIR) (per 1,000 mosquitoes) for *Cx. tarsalis* was higher and more consistent across counties, while the WNV PIR for *Cx. pipiens* was zero in many of the sampled counties (Figure 2C and 2D).

### Mosquito-Borne Diseases: Human Cases

The spatial distribution of average annual cumulative incidence (per 100,000 people) of WNV from 2012-2024 (Figure 2E and 2F) showed clustering of counties with higher cumulative incidences in western Nebraska compared to the rest of the state. This trend coincides with higher *Cx. tarsalis* abundance and consistently higher pooled infection rates compared to the rest of the state. A Global Moran’s I spatial autocorrelation test was used to assess the clustering pattern of the average annual cumulative incidence of human WNV cases from 2012-2024 at the county level (Figure 2E). There was a less than 1% chance that the observed clustering pattern of human WNV cases was the result of random chance (z-score = 3.66, p-value < 0.01), indicating significant spatial clustering of WNV cases.

### Mosquito Surveillance and Mosquito-Borne Diseases: Association Between Abundance, Infection Rates, and Human Cases

Nebraska counties with mosquito abundance, mosquito pooled infection rates, and human WNV cases were analyzed to determine if vector abundance and/or pooled infection rates were associated with human cases. In our preliminary analysis of all data from 2012-2024, we found that the data were not normally distributed and that overdispersion was present in all predictor and outcome variables. No heteroscedasticity was detected. In our negative binomial model using all data from 2012-2024, *Cx. pipiens* and *Cx. tarsalis* abundance and pooled infection rates were not statistically significant predictors (p-value > 0.05) of human WNV case counts.

We then conducted a separate analysis on a subset of that data to achieve a more contemporary perspective on counties reporting mosquito abundance, mosquito pooled infection rates, and WNV human case data for at least three of the four years from 2021-2024. The data in this subset was also not normally distributed. Overdispersion was present in all predictor and outcome variables, and no heteroscedasticity was detected. The gamma model with log link for our contemporary data subset (2021-2024) focusing on the outcome of WNV cumulative incidence per 100,000 also found no statistical significance among the same group of predictor variables.

Our negative binomial model for the contemporary data subset (2021-2024) indicated that the *Cx. Pipiens* pooled infection rate was significantly associated with total human WNV case counts (p-value <0.05). However, the range for the confidence interval for this predictor was below 1.0, indicating a likely negative relationship between *Cx. pipiens* pooled infection rates and total human case counts of WNV (Figure 3). Other predictors, including the *Cx. tarsalis* pooled infection rate and abundance for both mosquito species were not statistically significant (p-value > 0.05).

**Figure 3.**
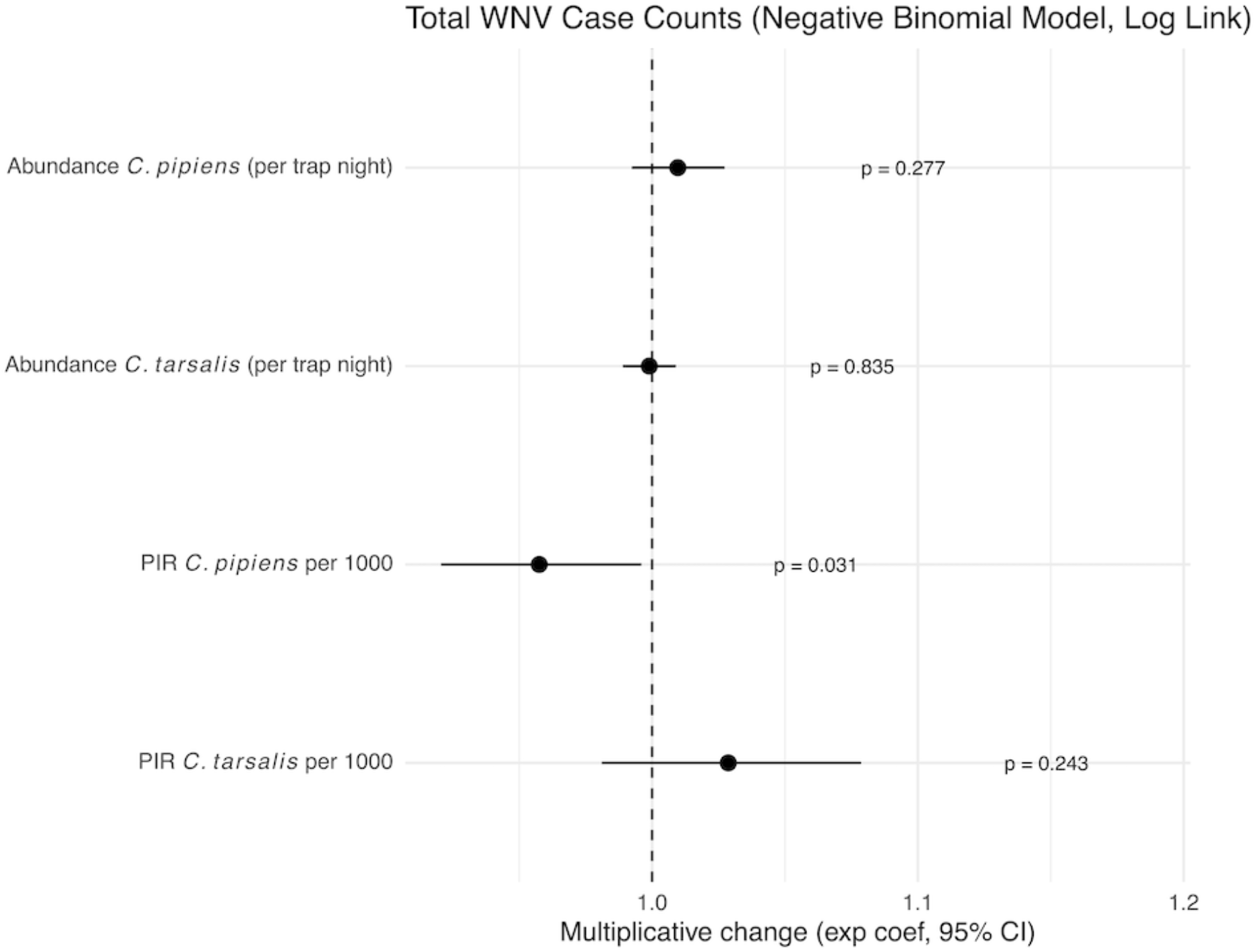
Coefficient plot for *Culex* species pooled infection rates and abundance with WNV human cases. A negative binomial model (log link) was used to evaluate associations between species-specific annual mosquito pooled infection rates and abundance per trap night with the outcome of total human WNV case counts across nine Nebraska counties from 2021-2024. Predictors include the pooled infection rates per 1,000 for *Cx. tarsalis* and *Cx. pipiens* and their respective annual abundances per trap night.

### Tick Surveillance: Tick Population Distribution, Abundance, and Pathogen Infection Rates

Populations of *A. americanum, D. variabilis*, and *I. scapularis* are established across Nebraska (Figure 4). However, the reported establishment of populations at the county level and average annual abundance per square meter differed between tick species. *A. americanum* populations were most concentrated in southeastern Nebraska (Figure 4B) while *D. variabilis* populations were reported across all sampled counties (Figure 4D). Conversely, *I. scapularis* populations were found only in two foci in eastern Nebraska, with the highest abundance in the northeastern region (Figure 4F).

**Figure 4.**
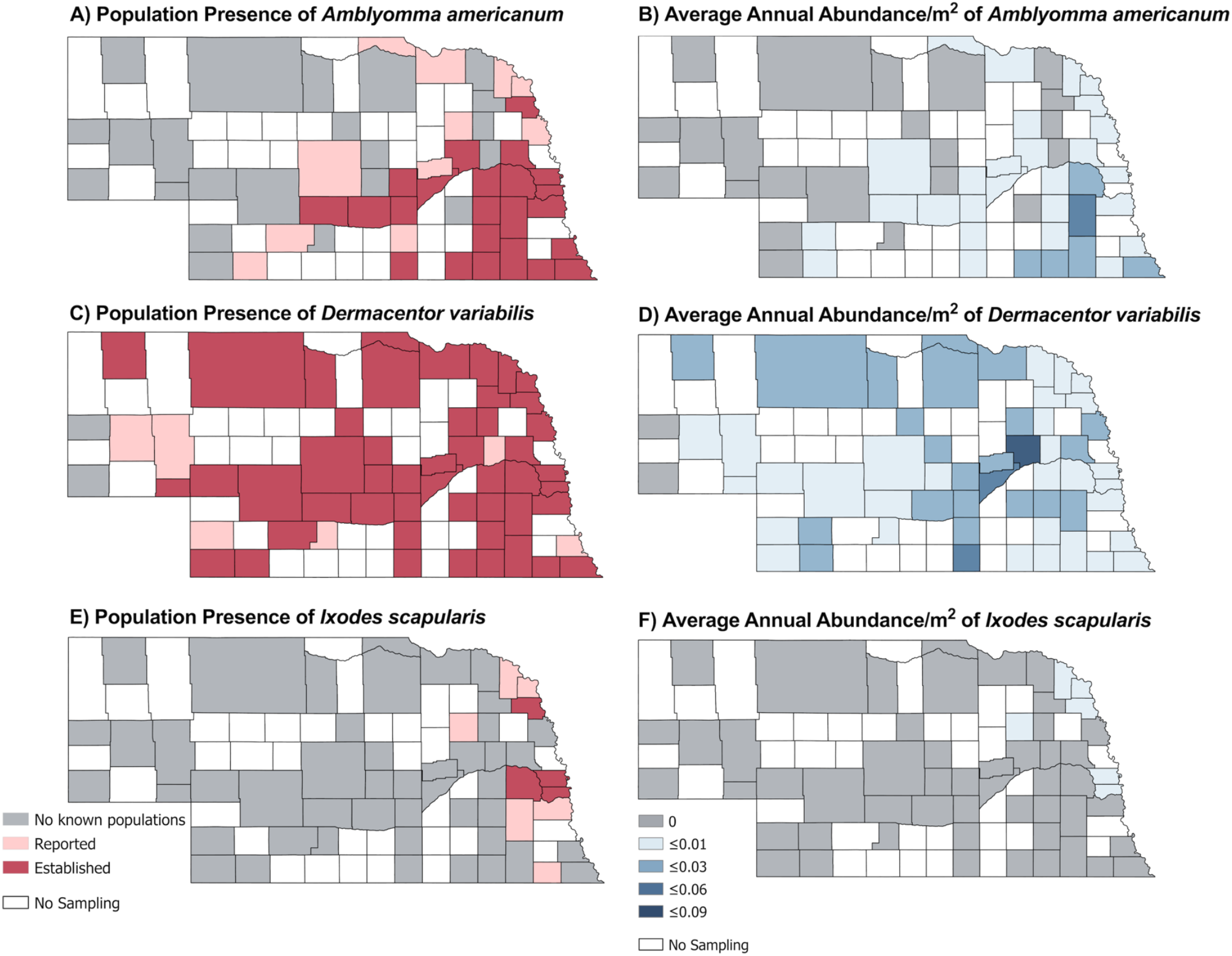
Comparison of tick species population reporting and abundance. A) Population presence of *A. americanum* in Nebraska and B) Average annual abundance per square meter from 2021-2024 of *A. americanum*. C) Population presence of *D. variabilis* in Nebraska and D) Average annual abundance per square meter from 2021-2024 of *D. variabilis*. E) Population presence of *I. scapularis* in Nebraska and F) Average annual abundance per square meter from 2021-2024 of *I. scapularis*.

Ticks collected during routine surveillance from 2021-2024 were tested for the presence of key tick-borne pathogens (Table 2). *Amblyomma americanum* ticks were found to be positive for *Ehrlichia chaffeensis* and *Ehrlichia ewingii. Dermacentor variabilis* ticks tested positive for *Ehrlichia ewingii, Francisella tularensis*, and *Rickettsia rickettsii. Anaplasma phagocytophilum, Borrelia miyamotoi*, and *Borrelia burgdorferi* were detected in *I. scapularis* ticks. Infection rates with these pathogens varied by tick species and location (Table 2).

### Tick-Borne Diseases: Human Cases

From 2021-2024, several Nebraska health department districts reported at least one case of anaplasmosis, ehrlichiosis, Lyme disease, rickettsiosis, or tularemia (Figure 5). Anaplasmosis cases were limited to the panhandle region of western Nebraska (Figure 5A), while ehrlichiosis cases were concentrated in southeastern Nebraska (Figure 5B). Diagnosed Lyme disease cases primarily occurred in eastern Nebraska (Figure 5C). Rickettsiosis cases were distributed sporadically across health department regions (Figure 5D) while tularemia cases were reported consistently for multiple years across most districts in the state (Figure 5E).

**Figure 5.**
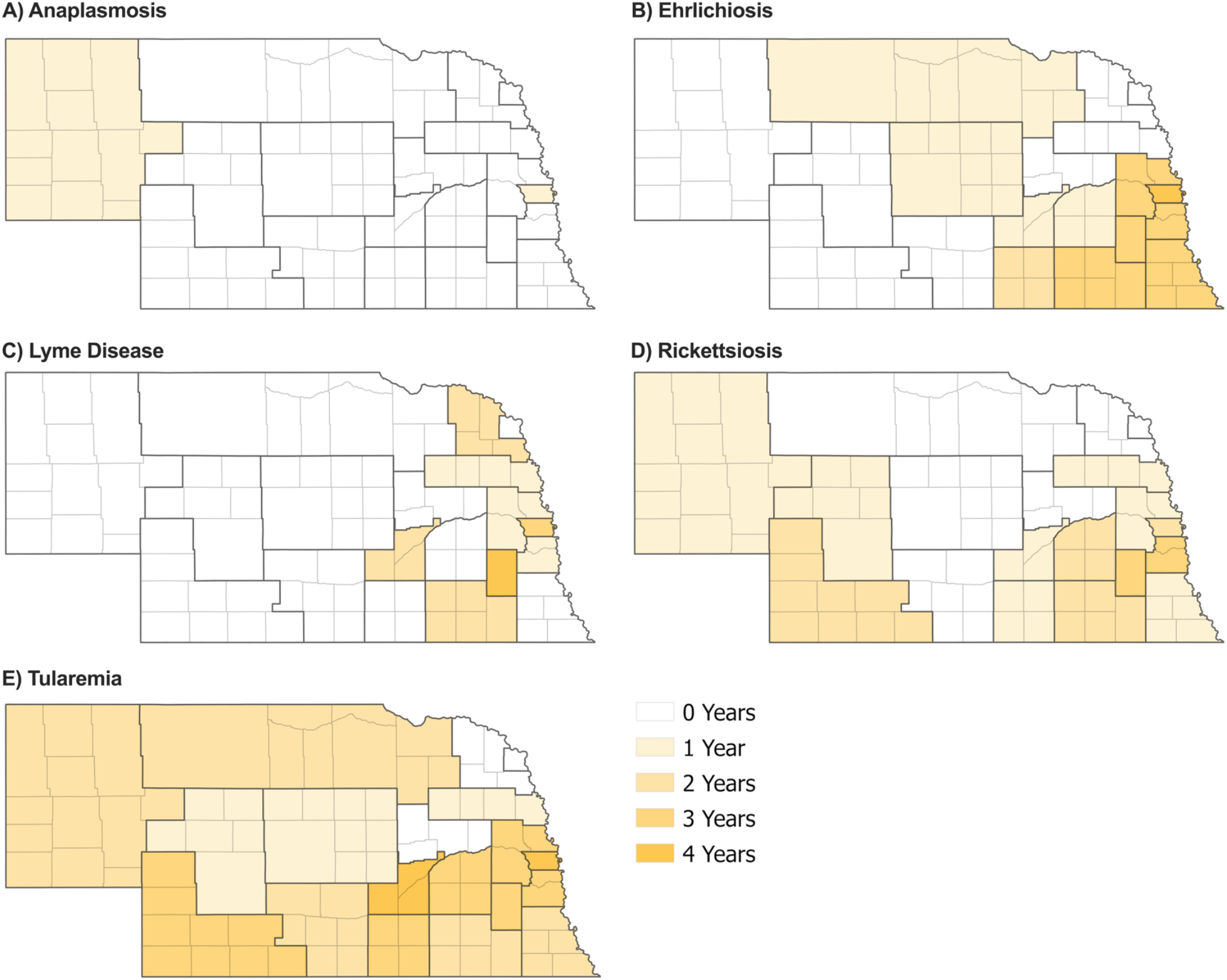
Number of years with reported anaplasmosis (A), ehrlichiosis (B), Lyme disease (C), rickettsiosis (D), and tularemia (E) human cases. The number of years (from 2021-2024) that each Nebraska health department reported at least one human case of anaplasmosis, ehrlichiosis, Lyme disease, rickettsiosis, or tularemia. Darker outlines represent health department regions and lighter outlines represent counties.

### Tick Surveillance and Tick-Borne Diseases: Association Between Tick Infection Rates and Human Cases

Health department regions with and without reported human cases of anaplasmosis, ehrlichiosis, Lyme disease, rickettsiosis, and tularemia were compared to determine if pathogen infection rates were associated with the presence of human cases. Infection rates were only calculated and compared in species of ticks that were tested for the presence of a specific pathogen. In general, pooled infection rates (per 1,000 ticks) were higher in health department regions reporting human disease for ehrlichiosis, Lyme disease, rickettsiosis, and tularemia (Figure 6). Median pooled tick infection rates for *Ehrlichia chaffeensis* or *Ehrlichia ewingii, Borrelia burgdorferi, Rickettsia* spp., and *Francisella tularensis* were elevated in these regions as well. Logistic regression analysis found no significant relationships between pooled infection rates in ticks and human cases reported at the health department level for all four diseases. A Wilcoxon rank-sum test indicated that higher *Ehrlichia spp*. infection rate in *A. americanum* ticks was associated with a higher likelihood of reporting the presence of a human case, but this relationship was not statistically significant. *A. phagocytophilum* detection in *I. scapularis* ticks was associated with health departments that did not report human cases of anaplasmosis (p-value <0.05; Figure 6).

**Figure 6.**
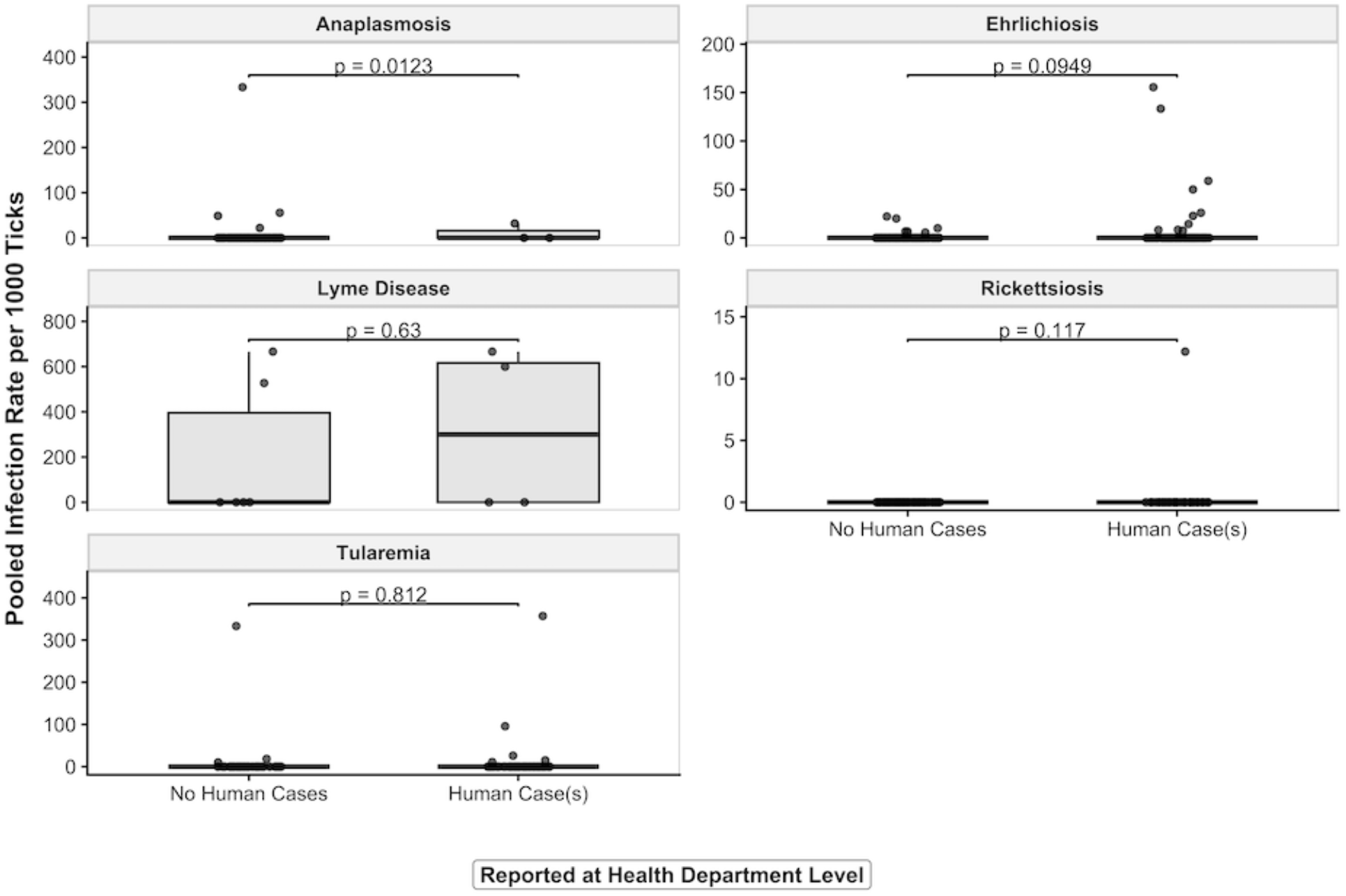
Pooled infection rates (per 1,000 ticks) for five tick-borne diseases by presence or absence of reported human cases at the health department level. Boxes represent interquartile ranges, horizontal lines indicate medians, and whiskers extend to 1.5x the interquartile range. Individual points show underlying observations. P-values are from Wilcoxon rank-sum tests. P-value <0.05 indicates significance.

## Discussion

### Mosquito Surveillance and Mosquito-Borne Diseases

The findings in this study specifically highlight the key role that WNV plays in Nebraska’s VBD landscape. While human cases of other arbovirus infections such as dengue fever and Zika virus infection have been reported, we limited our analysis to WNV because it is the most epidemiologically relevant arbovirus in Nebraska and the only arbovirus known to be consistently and autochthonously transmitted in the state. Retrospective surveillance data shows that increased WNV pooled infection rates among *Culex* mosquito populations correspond with increased reports of human WNV disease cases at the county level. This link supports the importance of entomological surveillance in determining long-term risk profiles at the county level. Despite WNV human cases being reported in nearly all 93 Nebraska counties, entomological surveillance is conducted in 40 counties. As of 2024-2025, only 22 counties are conducting mosquito surveillance, consistent with a larger trend in reductions of VBD surveillance across the US (Dye-Braumuller et al. 2022). Expanding entomological surveillance to include additional counties will allow for more comparisons between WNV mosquito pooled infection rates and human cases and better inform risk of WNV across the state.

The contemporary data subset (2021-2024) negative binomial model with log link for the outcome of WNV human case counts indicates that *Cx. pipiens* pooled infection rate is the only significant predictor. Specifically, higher *Cx. pipiens* pooled infection rates were associated with lower WNV human case counts, as the confidence interval for this predictor variable was below 1.0, suggesting a possible negative relationship. This observed relationship may reflect more complex ecological or mosquito behavior factors, such as trap sampling bias, rather than a true negative effect. It has been established that *Cx. pipiens* group mosquitoes drive more enzootic WNV amplification whereas *Cx. tarsalis* feeds on humans more frequently and may be more likely to transmit WNV in humans (Dunphy et al. 2019). *Culex tarsalis*, the primary WNV vector impacting humans in Nebraska, showed no statistically significant association between pooled infection rate and total human cases. Additionally, annual abundance per trap night measures for *Cx. pipiens* and *Cx. tarsalis* were not significant predictors of WNV human cases at the county level. This model based on our contemporary data subset likely also reflects trap and sampling bias, though the results of trapping could suggest that species-specific infection rates may have more complex relationships with human cases of WNV in Nebraska. Of the surveillance data that is currently available, both entomological and human case data likely underestimate WNV risk in Nebraska. Expanding mosquito surveillance efforts and improving case ascertainment would enable us to better understand the true relationship between entomological trends and human WNV risk in Nebraska.

### Tick Surveillance and Tick-Borne Diseases

The overall risk of TBDs in Nebraska is lower than MBDs; however, there is still risk for TBDs in Nebraska (Figure 1, Table 1). *Dermacentor variabilis* dominated tick collections throughout the state, with detections of *A. americanum* in the southeastern region and minimal collections yielding *I. scapularis* ticks in the northeast. This adds to previous literature where historical examination of tick submissions in Nebraska from 1911 – 2011 noted that *D. variabilis* (American dog tick) was the most common tick identified while *A. americanum* (lone star tick) was first observed in 1944 (Cortinas and Spomer 2014). However, consistent *A. americanum* observations did not start until the late 1980s in the most southeast part of the state (Cortinas and Spomer 2013, 2014). The first established *I. scapularis* (blacklegged tick) populations in Nebraska were reported in 2019 in the east-central region, with the first detection of *Borrelia burgdorferi* in these ticks associated with local Lyme disease transmission reported in 2021 (Nielsen et al. 2020, Hamik et al. 2023). Lyme disease reports were included in this study as diagnosed human cases are reported to Nebraska public health districts. However, autochthonous transmission of *B. burgdorferi* has only been definitively established in Thurston County (Nielsen et al. 2020, Hamik et al. 2023). The limited presence of *I. scapularis* populations in the state is consistent with this pattern. While there were a notable number of Lyme disease cases reported during this timeframe, the data were not suggestive of widespread local transmission. The findings in this study emphasize that Nebraska’s tick populations support the persistence of several other pathogens capable of maintaining localized enzootic cycles. The recurrence of tick pools testing positive for pathogens known to cause anaplasmosis, ehrlichiosis, rickettsiosis, and tularemia points to local transmission. It is also important to note that while human cases of tularemia are most often related to tick bites, the handling of infected small mammals, such as rabbits, can also lead to transmission (Sharma et al. 2023). Furthermore, while mosquitoes are more mobile, exposure to ticks and the subsequent risk of tick-borne pathogens depends on human-tick encounters, leading to varying risk (Fischhoff et al. 2019). The persistent presence of disease-causing pathogens in tick populations and possible different modes of transmission, supports continued tick surveillance and expanded geographic coverage to detect shifts in vector and pathogen distributions.

### Strengths and Limitations: Overall

Strengths of this study include the statewide integration of entomological surveillance and human case data, allowing for robust analyses between vector pooled infection rates and human disease. This comprehensive approach provides a stronger foundation for predictive modeling and informed public health interventions.

This study was limited by the underreporting of human VBD cases, inconsistent sampling of mosquito and tick populations across the state, and reliance on trapping methods that may influence species-specific catch rates. Specifically, Nebraska utilizes CDC light traps for mosquito collections. While *Cx. tarsalis* is reliably collected with CDC light traps, gravid traps are more effective at collecting *Cx. pipiens* (Reiter et al. 1986, Boze et al. 2021, CDC 2024f). Additionally, short surveillance windows for tick sampling and low detection numbers for rarer pathogens (particularly among *Ixodes* ticks) limit our ability to draw conclusions about long-term TBD trends.

### Strengths and Limitations: Mosquito Surveillance and Mosquito-Borne Diseases

Our modeling approach used with the WNV entomological and human case data had several limitations. The contemporary dataset included only nine counties over four years. We were therefore limited in our statistical power, stability of parameter estimates, and the ability to accurately detect non-normality, heteroscedasticity, or overdispersion with standard tests. Additionally, annual aggregation of mosquito and human case data may obscure intra-seasonal dynamics and may not account for temporal changes between vector WNV infections and human cases. Additionally, reported human cases depend on diagnoses, access to diagnostic tests, and possible healthcare-seeking behaviors which may vary across different populations. Our chosen model also relied on distribution assumptions that can be difficult to verify with small sample sizes. While indicated for the small sample sizes, treating county and year as fixed effects further reduces generalizability. Other environmental factors known to influence WNV transmission were also not modeled, limiting causal interpretations.

### Strengths and Limitations: Tick Surveillance and Tick-Borne Diseases

Since TBD data is reported at the health department level and case counts below six are suppressed, our analyses could not be performed at the county level, thus reducing geographic data resolution and interpretations at fine scales. Tick surveillance was conducted at the county level, requiring extrapolation of entomological data to health department boundaries, which likely introduced bias in the associations. Tick abundance and infection rates also relied on sampling intensity and may not accurately reflect true tick population dynamics or infections across all counties. Also, comparisons with publicly available tick species population reporting were qualitative and may have been influenced by differences in surveillance rather than variation in tick presence alone. Statistical power was limited due to low numbers of human cases across TBDs, possibly contributing to the lack of significant associations in our Wilcoxon rank-sum tests.

### Conclusions and Future Directions

In this study, trends in vector abundance, vector infection rates, and human cases to determine the association between vector prevalence, infection rates, and human VBD risk were assessed. The findings of this study reinforce the need to expand integrated vector surveillance and human VBD case reporting programs across Nebraska. Mosquito WNV pooled infection rates, particularly in *Cx. tarsalis*, serve as valuable and actionable indicators for potential human cases, as demonstrated in Iowa, Nebraska, and Colorado (Schweitzer et al. 2006; Bolling et al. 2009; Fauver et al. 2016; Dunphy et al. 2019). Additionally, because mosquito trapping occurs every other week during a given season, there is the potential to identify key early warning signals of elevated WNV risk. Broadening current surveillance efforts and strengthening tick monitoring will be essential to understand the risk of VBD transmission in the state as environmental conditions and vector distributions continue to shift over time.

Nebraska’s current VBD burden is driven primarily by WNV and emerging tick-borne pathogens. Continued and expanded investment in integrated surveillance, linking vector, pathogen, and human data, will enhance early warning systems, guide targeted interventions, and ultimately strengthen the state’s capacity to mitigate both established and emerging VBD threats.

## Data Availability

All data produced in the present study are available upon reasonable request to the authors.

https://www.cdc.gov/west-nile-virus/data-maps/historic-data.html

https://dhhs.ne.gov/Pages/Vector-Borne-Disease-Data-and-Statistics.aspx

https://dhhs.ne.gov/Pages/Tick-Borne-Disease-Data-and-Statistics.aspx

https://dhhs.ne.gov/Pages/West-Nile-Virus-Data.aspx

## Acknowledgements

We would like to acknowledge the national arbovirus surveillance system, ArboNET, for providing human WNV case data for the state of Nebraska. We would also like to acknowledge Nebraska DHHS for providing human tick-borne disease case data and tick surveillance data and the NPHL and DHHS for supporting mosquito collections.

